# Semantic and phonetic markers in schizophrenia-spectrum disorders; a combinatory machine learning approach

**DOI:** 10.1101/2022.07.13.22277577

**Authors:** A.E. Voppel, J.N. de Boer, S.G. Brederoo, H.G. Schnack, I.E.C. Sommer

**Affiliations:** UMCG, Universiteit Groningen; UMC Utrecht, Universiteit Utrecht; Universiteit Utrecht

## Abstract

**Introduction:** Speech is a promising marker for schizophrenia-spectrum disorder diagnosis, as it closely reflects symptoms. Previous approaches have made use of different feature domains of speech in classification, including semantic and phonetic features. However, an examination of the relative contribution and accuracy per domain remains an area of active investigation. Here, we examine these domains (i.e. phonetic and semantic) separately and in combination.

**Methods:** Using a semi-structured interview with neutral topics, speech of 94 schizophrenia-spectrum subjects (SSD) and 73 healthy controls (HC) was recorded. Phonetic features were extracted using a standardized feature set, and transcribed interviews were used to assess word connectedness using a word2vec model. Separate cross-validated random forest classifiers were trained on each feature domain. A third, combinatory classifier was used to combine features from both domains.

**Results:** The phonetic domain random forest achieved 81% accuracy in classifying SSD from HC. For the semantic domain, the classifier reached an accuracy of 80% with a sparse set of features with 10-fold cross-validation. Joining features from the domains, the combined classifier reached 85% accuracy, significantly improving on models trained on separate domains. Top features were fragmented speech for phonetic and variance of connectedness for semantic, with both being the top features for the combined classifier.

**Discussion:** Both semantic and phonetic domains achieved similar results compared with previous research. Combining these features shows the relative value of each domain, as well as the increased classification performance from implementing features from multiple domains. Explainability of models and their feature importance is a requirement for future clinical applications.

## Introduction

Recently, computational speech analysis has become a promising marker for schizophrenia-spectrum disorders (SSD) (Corcoran et al., 2020; de Boer, Brederoo, et al., 2020; Parola et al., 2020). Natural language processing (NLP) has been used to predict conversion to psychosis in ultra-high-risk individuals (Bedi et al., 2015; Corcoran et al., 2018; Spencer et al., 2021), classify healthy controls from subjects with schizophrenia-spectrum disorders (Bar et al., 2019; Elvevåg et al., 2010; Voppel et al., 2021) and differentiate patients with two different diagnoses (Mota et al., 2014). Quantified speech features calculated using NLP methods have been related to specific symptoms such as formal thought disorder (Elvevåg et al., 2007; Mackinley et al., 2020) as well as categories such as negative symptoms (Cohen et al., 2020; Compton et al., 2018). Here, we investigate the relative strengths of different speech characteristics and their combination in differentiating people with SSD from healthy controls.

Researchers have made use of different domains of speech for these investigations, including phonetic, semantic and other linguistic features with results in the 80-100% accuracy range. Phonetic sources of information include pausation, speech rate, intonation and percentage of spoken time (de Boer et al., 2021; for a review see Parola et al., 2020a). Symptoms commonly associated with the phonetic or acoustic domain include alogia (poverty of speech), blunted affect and other negative symptoms, as well as some positive symptoms such as pressured speech (Alpert et al., 2002). Semantic features include discourse coherence, semantic density and connectedness in language (Corcoran et al., 2018; Elvevåg et al., 2007; Rezaii et al., 2019; Voppel et al., 2021). These approaches make use of computational models of semantic information present in language (de Boer et al., 2018). Symptoms related to these methods are mostly positive symptoms including formal thought disorder, tangentiality and incoherence of speech (Bedi et al., 2013; Mackinley et al., 2020; Tang et al., 2021).

Rapidly developing NLP techniques have resulted in a multitude of features. As an example, Marmar and colleagues made use of tens of thousands of acoustic features in classifying post-traumatic stress syndrome (Marmar et al., 2019). Because of the large number of possible features, interpretation and comparability of findings is problematic (de Boer, Brederoo, et al., 2020). Explainability of features and algorithms is a critical step towards implementing machine learning models in clinical practice (Chandler et al., 2020; Tonekaboni et al., 2019). The relative added value of features and indeed of combining the phonetic and semantic domains has remained obfuscated. Heterogeneity of SSD further increases the problem of classification approaches, as two subjects with SSD can have zero overlapping symptoms and thus might not be detectable by using a single domain of speech features for classification. The problem of this heterogeneity can be tackled by employing multiple sets of features, reflecting a larger set of speech characteristics.

In this study, we train separate classifiers for both the phonetic and semantic domain. We investigate their performance and examine top features, then merge the features in a combined classifier to assess the value of using a combinatory approach. We further delve into the relative strengths and weaknesses of the phonetic and the semantic domain in classifying schizophrenia-spectrum disorders, a step towards usage of speech features in clinical practice.

## Methods

### Participants

Language was recorded from a semi-structured interview of 94 participants with SSD and 73 healthy controls, adding up to 167 participants. All patients were diagnosed with schizophrenia, psychosis not otherwise specificied (NOS), schizophreniform or schizoaffective disorder by the treating physician, and confirmed using either the CASH or the MINI diagnostic interviews (Andreasen et al., 1992; Sheehan et al., 1998). Inclusion criteria for healthy controls were the absence of a psychiatric diagnosis and history, with the exception of depression or anxiety disorders in full remission. All participants were 18 or above and native Dutch speakers. Participants were informed that the interview was analyzed for ‘general experiences’ to prevent participants focusing on their speech or pronunciation. After completion, participants were told that the research investigated their speech. Before enrollment, all participants gave written informed consent. The study was approved by the University Medical Center Utrecht ethical review board. Antipsychotic medication use was calculated as chlorpromazine equivalents in milligram per day following Leucht (2014). Symptoms were assessed using the positive and negative syndrome scale (PANSS -Kay et al., 1987).

### Interview procedure

Speech was recorded using a digital TASCAM DR-40 recording device using head-worn AKG-C544l cardioid microphones, using a separate recording channel for participant and interviewer with a sampling rate of 44.1 kHz. The semi-structured interview was performed by trained interviewers; for an elaborate description of the interview methodology, see previous reports by our group (de Boer, van Hoogdalem, et al., 2020; de Boer, Voppel, et al., 2020; Voppel et al., 2021). Topics discussed in the interview were neutral, avoiding specific illness related topics, and participants could skip questions if they wanted. For a list of questions, see supplemental materials of de Boer, Voppel et al., 2021.

### Phonetic domain: processing and parameters

To remove crosstalk the following steps were taken: 1) the ‘annotate silences’ function in PRAAT (Boersma & Weenink, 2013) was used on the interviewer’s channel; 2) all resulting speech segments in which the interviewer was silent were selected on the participants channel; 3) the resulting speech segments were concatenated to a new audio file containing only segments of the participants’ speech. Using openSMILE (Eyben et al., 2013), the extended GeMAPS parameter set was used to extract a total of 88 parameters at the speaker level. The parameters can be divided into 6 temporal parameters such as speech rate, 24 frequency parameters, 43 spectral parameters such as Mel-frequency cepstral coefficients, and 14 energy/amplitude parameters such as intensity. This procedure is the same as previous research done by our group (de Boer et al., 2021). Select features derived using the parameter set can be characterized as acoustic, notably pauses and speech rate, instead of phonetic as they are only indirectly related to speech produced. Here we have chosen to categorize all features from this feature set as phonetic.

### Semantic domain: processing and parameters

Speech was transcribed according to the CLAN-CHILDES protocol (MacWhinney, 2000). Following transcription, the text was vectorized using a word2vec language model (Mikolov et al., 2013). Following previous research from our group (Voppel et al., 2021), a moving window approach was employed to calculate word-to-word similarity within windows of words sized 5-10. Within a window, the similarity of each word to each other word within that window was computed and then averaged, resulting in a single word connectedness in the window. The window then moved one position further, a new connectedness value was computed, and this procedure was repeated until the end of the transcript, resulting in a series of word connectedness values. From this series of connectedness values, variance, mean, and minimum of connectedness between windows 5 and 10 was computed per subject.

### Random forest classifier models

Separate random forest classifiers were trained to assess feature accuracy per domain. Models were trained in R, using the caret software package (Kuhn, 2008; R Core Team & others, 2013). The models used 10-fold cross-validation, where 90% of the data set is used as training with a randomly chosen 10% as a testing sample, repeated ten times until all samples have served as a testing sample. 500 trees were grown, with number of features sampled per decision split the square root of the total number of features. To combine predicting features from different domains a third model was trained. The features of both the phonetic and semantic domains were used as input in this final random forest model. This model was then once again cross-validated, using 10-fold cross-validation.

Probability estimates for each of the trained models (phonetic, semantic and combined) were used to generate receiver operator curves (ROC) and areas under the curve (AUC). From each of the trained classifiers, predictor feature importance (Gini-importance score) was calculated, measuring how much worse the model becomes when replacing each predictor in decision trees with random data distributed according to the SSD:HC ratio. The difference between the original model performance and performance without the feature is then taken as the added value of the feature.

### Statistics

Statistical analysis was performed in R (R Core Team & others, 2013). Demographic characteristics were compared between the groups using ANOVAs for continuous variables, and χ2 tests were used for categorical variables. Pearson’s correlational analyses were performed between continuous variables as possible confounders. To assess relative model performance significance, we used McNemar’s test (McNemar, 1947).

## Results

The 94 participants with SSD and 73 healthy controls did not differ significantly in age, sex, see Table 1. Healthy controls had received significantly more education than SSD patients, *p*=0.003, which was not unexpected, as SSD commonly develops during the educational years; we therefore investigated the relation between education and the top informative classifier features. see table 3. Groups did not differ in parental educational levels.

**Table 1.**
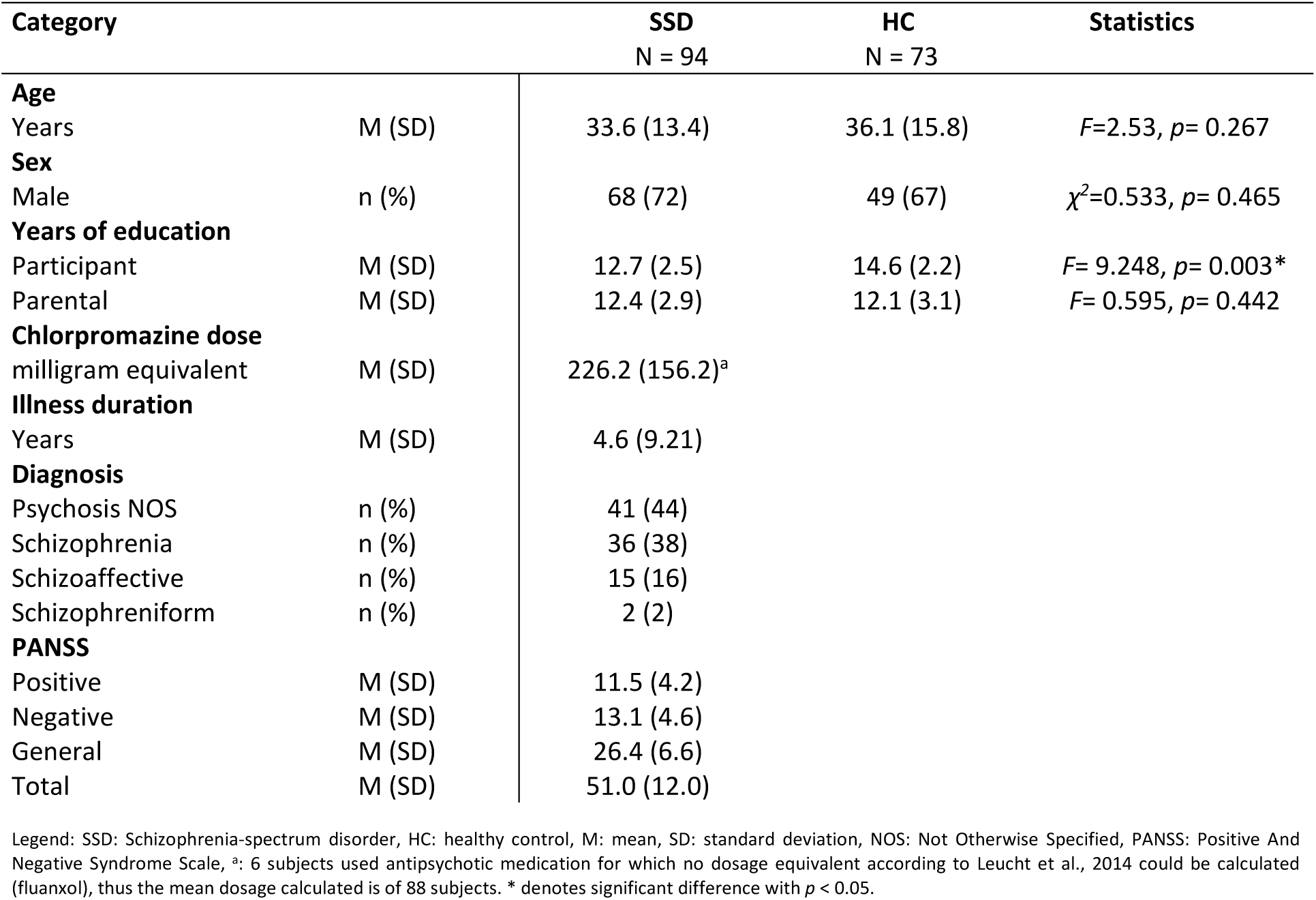
Demographics

| Category |  | SSD<br>N = 94 | HC<br>N = 73 | Statistics |
| --- | --- | --- | --- | --- |
| **Age** |  |  |  |  |
| Years | M (SD) | 33.6 (13.4) | 36.1 (15.8) | $F=2.53, p= 0.267$ |
| **Sex** |  |  |  |  |
| Male | n (%) | 68 (72) | 49 (67) | $\chi^2=0.533, p= 0.465$ |
| **Years of education** |  |  |  |  |
| Participant | M (SD) | 12.7 (2.5) | 14.6 (2.2) | $F= 9.248, p= 0.003^*$ |
| Parental | M (SD) | 12.4 (2.9) | 12.1 (3.1) | $F= 0.595, p= 0.442$ |
| **Chlorpromazine dose** |  |  |  |  |
| milligram equivalent | M (SD) | 226.2 (156.2)<sup>a</sup> |  |  |
| **Illness duration** |  |  |  |  |
| Years | M (SD) | 4.6 (9.21) |  |  |
| **Diagnosis** |  |  |  |  |
| Psychosis NOS | n (%) | 41 (44) |  |  |
| Schizophrenia | n (%) | 36 (38) |  |  |
| Schizoaffective | n (%) | 15 (16) |  |  |
| Schizophreniform | n (%) | 2 (2) |  |  |
| **PANSS** |  |  |  |  |
| Positive | M (SD) | 11.5 (4.2) |  |  |
| Negative | M (SD) | 13.1 (4.6) |  |  |
| General | M (SD) | 26.4 (6.6) |  |  |
| Total | M (SD) | 51.0 (12.0) |  |  |

Legend: SSD: Schizophrenia-spectrum disorder, HC: healthy control, M: mean, SD: standard deviation, NOS: Not Otherwise Specified, PANSS: Positive And Negative Syndrome Scale, <sup>a</sup>: 6 subjects used antipsychotic medication for which no dosage equivalent according to Leucht et al., 2014 could be calculated (fluanxol), thus the mean dosage calculated is of 88 subjects. \* denotes significant difference with $p < 0.05$ .

**Table 2.** Classifier performance for phonetic, semantic and combined models. Bold denotes best scoring model per measure. Legend: Sens.; sensitivity, spec.: specificity, AUC: Area under the curve, CI: confidence interval.

|  | Accuracy | Sens. (95% CI) | Spec. (95% CI) | AUC (95% CI) |
| --- | --- | --- | --- | --- |
| Phonetic | 0.81 | 0.89 (0.82-0.94) | 0.70 (0.59-0.79) | 0.82 (0.76-0.88) |
| Semantic | 0.80 | 0.81 (0.72-0.88) | **0.78** (0.67-0.86) | 0.83 (0.77-0.89) |
| Combined | **0.85** | **0.92** (0.84-0.96) | 0.77 (0.66-0.85) | **0.88** (0.83-0.93) |

**Table 3.** Top informative features of combined classifier

| Feature name | Reflecting | SSD | HC | ANOVA | | Correlation with YOE<br>Pearson's $r$ |
| --- | --- | --- | --- | --- | --- | --- |
| | | mean (sd) | mean (sd) | $F$ | $p$ | |
| VoicedSegmentsPerSec | Average number of continuous voiced regions per second (more segments indicates more fragmented speech with short speech segments). | 1.651<br>(0.451) | 1.326<br>(0.212) | 39.009 | < .001** | -.247** |
| var_simple_7 | Variance of word connectedness, window size 7 | 0.00622<br>(0.000768) | 0.00567<br>(0.000311) | 22.126 | < .001** | n.s. |
| slopeV5001500_sma3nz_amean | Mean spectral Slope 0-500 Hz and 500-1500 Hz, reflecting tension | -0.0229<br>(0.00335) | -0.0248<br>(0.00301) | 2.168 | 0.143 | -.191* |
| var_simple_10 | Variance of word connectedness, window size 10 | 0.00622<br>(0.000713) | 0.00571<br>(0.000293) | 24.811 | < .001** | n.s. |
| var_simple_8 | Variance of word connectedness, window size 8 | 0.00622<br>(0.000758) | 0.00570<br>(0.000296) | 23.228 | < .001** | n.s. |

Legend: SSD: schizophrenia-spectrum disorders; HC: healthy controls; sd: standard deviation; YOE: Years of educations; n.s.: not significant; \* denotes significance at $p < 0.05$ ; \*\* denotes significance at $p < 0.01$

### Phonetic classifier

The 10-fold cross-validated random-forest classifier using features from the phonetic domain had an accuracy of 81%, with a sensitivity of 89%, and specificity 70%, see Table 2. The AUC-ROC was 0.82 in classifying subjects with SSD from HC. The top feature as ranked by Gini importance was voiced segments per second, reflecting shorter, fragmented speech in the SSD group, see table 3. Top 10 features are ranked in figure 1a.

**Figure 1.**
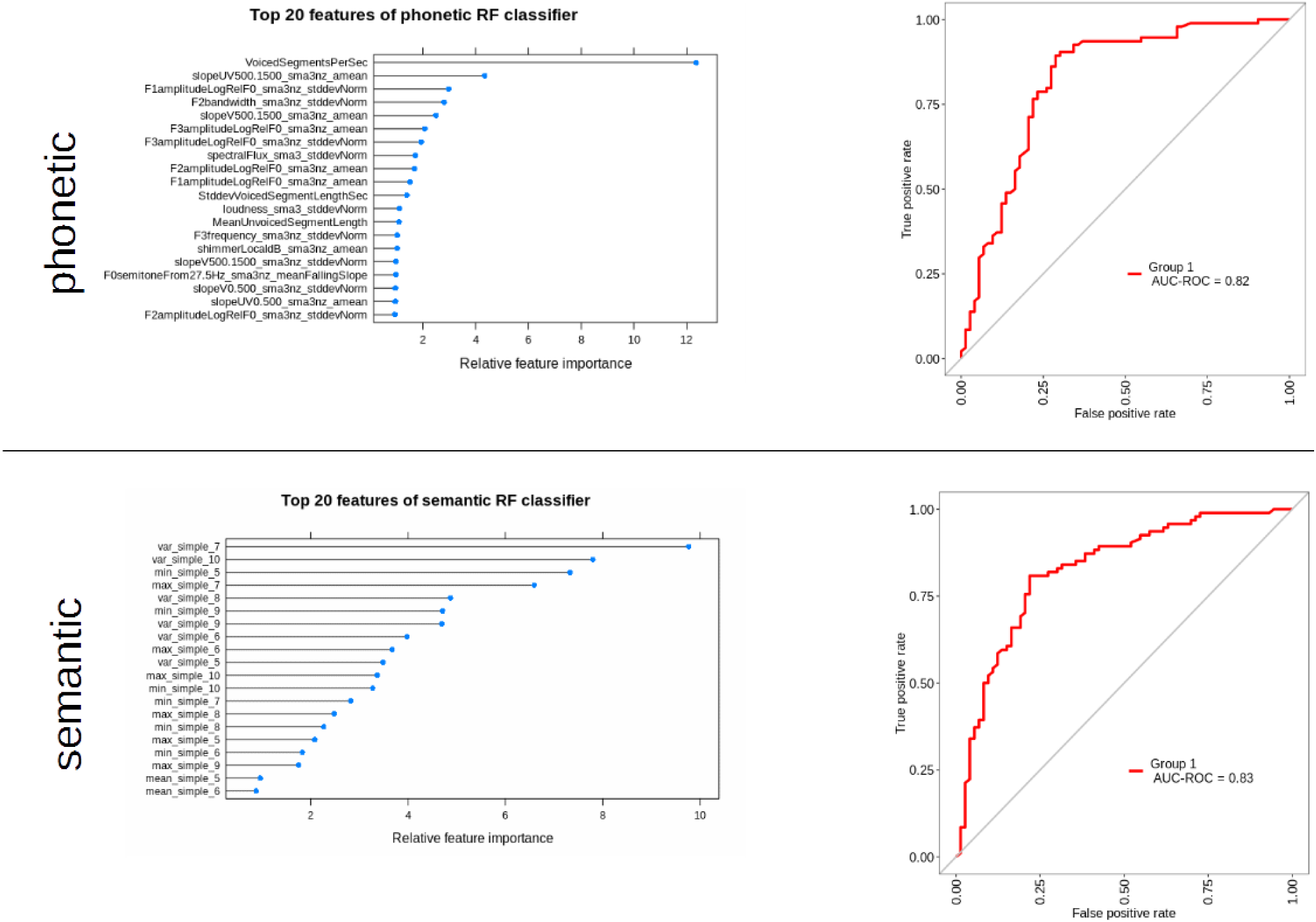
Separate domain classifier performance and features. **Top:** Phonetic domain classifier performance. Top informative features of the phonetic classifier, left, ranked by Gini feature importance. Right, AUC-ROC curve of the phonetic classifier. **Bottom:** Semantic domain classifier performance, with top informative features left and AUC-ROC curve of semantic classifier right.

### Semantic classifier

For features from the semantic domain, the 10-fold cross-validated random forest reached an accuracy of 80%. Sensitivity of the model was 81%, and specificity 78%. The AUC-ROC using semantic features was 0.83, with the top feature being variance of connectedness in a window of 7, indicating an increase in variance of sentence-level word connectedness in SSD compared to HC, see table 3. Gini ranking of top 10 features for the semantic domain are shown in figure 1b.

### Combined classifier

The combined classifier, trained with features from both domains, reached an accuracy of 85%, with sensitivity reaching 92% and specificity 77%. The AUC-ROC reached 0.88. The top informative feature was the top phonetic feature voiced segments per second, with the 2^nd^ most informative feature variance of connectedness in a window of 7, the highest ranked feature for the semantic classifier; for a full list of features, see figure 2.

**Figure 2.**
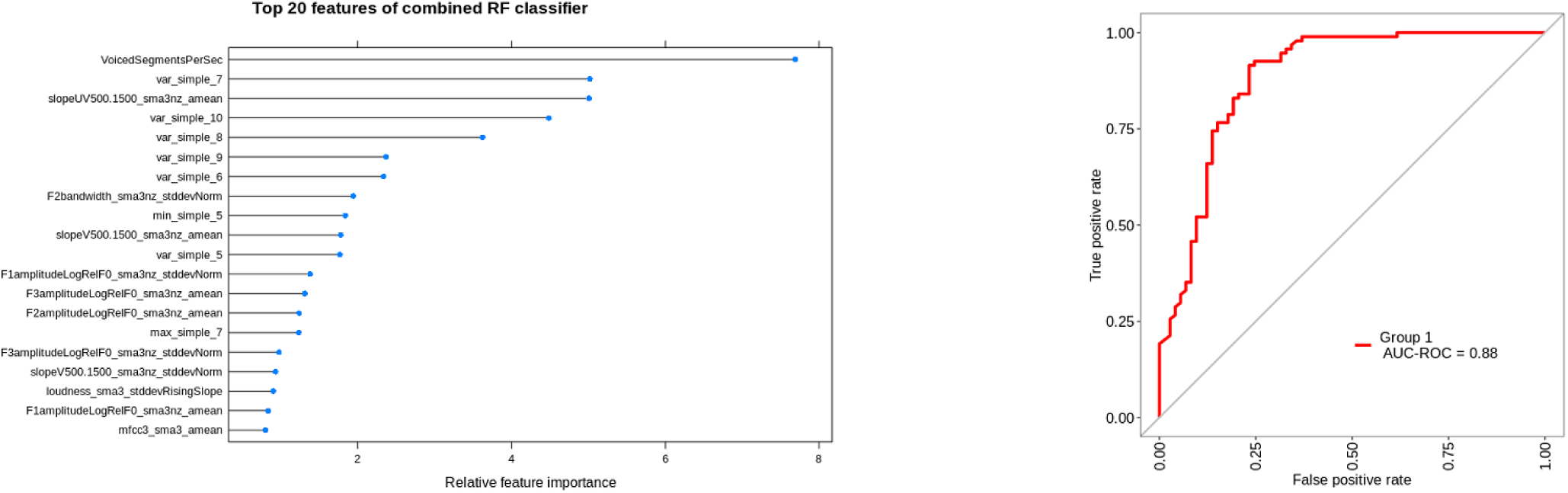
Combined domains classifier Left; top features of random forest classifier, ranked by Gini coefficient. Right, AUC-ROC curve of the combined classifier.

### Comparing classifier performance and misclassifications

Classification results of phonetic, semantic and combinatory classifiers are shown in table 2. Statistically comparing classification model performance using McNemar’s test (McNemar, 1947) showed the combined domain classifier significantly better compared to the phonetic classifier, McNemar’s test *χ*^*2*^ = 4.800, *p* = 0.0285, as was the combined domain classifier compared to the semantic classifier, *χ*^*2*^ = 6.125, *p* = 0.0133.

To evaluate misclassifications, we compared overlap in misclassifications per modality and in the combined model. For healthy controls, 24% of subjects misclassified by the phonetic domain classifier were also misclassified by the combined classifier; the overlap in misclassification was 33% for semantic and combined classifiers. In total, 8 healthy controls were misclassified by all three models.

28% of SSD subjects misclassified in the phonetic classifier were also misclassified by the combined model, with the same 28% overlap between the semantic classifier and the combined classifier SSD misclassifications. However, only a single SSD subject was misclassified by all 3 models, showing the added sensitivity by combining different sources of information.

## Discussion

This study shows that speech features of phonetic and semantic domains can be combined to improve accuracy of speech classifiers for SSD. The two highest-ranked features in the combined classifier were the two top features in the domain-specific classifiers, showing the added information in combining domains to improve classifier performance. Classifiers trained on phonetic and semantic features separately were comparable in overall accuracy, with some relative strengths for sensitivity (phonetic domain) and specificity (semantic). The classifier combining feature domains performs significantly better than the separate models on overall performance, accuracy, as well as sensitivity, and achieved 1% lower specificity than the semantic domain classifier.

In training separate classifiers for both the phonetic and semantic domain, we replicate previous findings showing their applicability in classifying subjects with schizophrenia-spectrum disorders from healthy controls (Compton et al., 2018; Corcoran et al., 2018; de Boer et al., 2021; Parola et al., 2020; Rezaii et al., 2019; Voppel et al., 2021). The most informative phonetic feature was supported by previous literature, as speech rate and pausing patterns are often disturbed in schizophrenia-spectrum disorders (Parola et al., 2020; see also table 3). For the semantic classifier we found that the most informative feature to be variance of word connectedness at window size 7, which was also reported previously research by our group in a smaller sample (Voppel et al. 2021). The window size of 7 reflects word connectedness at sentence level, which consistently outperforms word-to-word connectedness in classification algorithms (Corcoran et al., 2018; de Boer et al., 2018).

There was a substantial difference in years of education between the groups. While no significant correlations were found between years of education and the semantic features used in this study, as expected based on previous findings (Voppel et al., 2021), we found significant correlations between years of education and the most informative phonetic features, shown in table 3. It is thus possible that years of education explains part of the phonetic feature importance, serving as a confounder. Some other limitations should be observed. The present study made use of cross-validation to estimate the generalizability of the models; while this a valid approach, the usage of an independent validation sample is the gold standard approach to test for possible overfitting on the training set. Smoking status of participants and controls was not available, but it is known to have an impact on aspects of speech such as frequency and voice quality (Gonzalez, 2003; Ma et al., 2021).

Our results show that, using features derived from previous findings, speech feature domains can be combined to reach greater accuracy. Importantly, we here show the relative features both per domain and in a combination, allowing us to retrieve the added value of features. Adding more features allows for more complex models that better handle heterogeneous group classification, but more complex models make it harder to explain results. Similarly, various machine learning algorithms can suffer from a lack of explainability, with “black box” models being undesired in the clinical application of machine learning models (Samek et al., 2017). Indeed, explainability of models and the importance of features therein can be a reason for choosing a slightly worse-performing classification algorithm over a more accurate one that lacks explainability (Chandler et al., 2020). Through assessing and choosing specific features, explainable and simple algorithms can be created for future clinical applications (Tonekaboni et al., 2019).

Future research could improve on the findings presented here. While we used a relatively sparse set of semantic features and a standard feature set for phonetic assessment of speech, a stricter selection of features would lead to better interpretability. Conversely, future research could derive additional features from other domains of language and speech, such as syntax, if these domains or features capture more heterogeneity. Caution should be taken to preserve explainability if such an approach is taken, by choosing for example a voting algorithm and/or a sparse set of explainable features.

While we focused here on classifying subjects, a similar combinatory methodology can also be used for different applications such as monitoring treatment or relapse prevention. In these cases, including other specific features relevant to the application. For example, to predict relapse one might include specific nonverbal features associated with negative symptoms (Tahir et al., 2019) while a researcher investigating the severity of memory impairments might want to incorporate increased pausing as a specific domain (Oomen et al., 2021).

The combination of reproducible, objective features spanning different domains derived from a single recording makes language and speech analysis a prime target as a marker for schizophrenia-spectrum disorder (Tan & Rossell, 2020). Once a transcription is available, characteristics like syntax, semantics, sentiment analysis and specific word usage can be combined with phonetic information, all with no added burden for participants.

In sum, we showed that phonetic and semantic features of speech can be combined to classify subjects with schizophrenia-spectrum disorders from healthy controls with 85% accuracy. While both domains of features can be used separately, the combination of domains performs better. An important consideration in the future development of speech classifiers remains their explainability, in preparation of a clinically acceptable tool to aid diagnosis or treatment monitoring.

## Data Availability

Anonymous data produced in the present study are available upon reasonable request to the authors

## Acknowledgements

We are grateful to all participants of the study, as well as to all research interns for their help with data collection and preparation.

